# Intersectional inequalities in demand satisfied by modern contraceptive methods among women in sub-Saharan Africa, 1994 to 2024: a multilevel analysis of individual heterogeneity and discriminatory accuracy

**DOI:** 10.64898/2026.11.12.26367922

**Authors:** Patrick Kaonga, Sylvia Masambi, Musopa Nkole, Alice Ngoma-Hazemba

## Abstract

**Background:** Demand for family planning satisfied by modern methods is Sustainable Development Goal (SDG) indicator 3.7.1, and countries have committed to universal access to family planning by 2030. Inequalities are usually reported one social dimension at a time. This hides how social characteristics combine and leaves open whether programmes should target social groups or strengthen services for all women. We measured intersectional inequalities in demand satisfied by modern methods among women in sub-Saharan Africa over three decades, assessed how well social position discriminates between women, and identified groups left behind.

**Methods and findings:** We did a preregistered cross-sectional analysis of 139 Demographic and Health Surveys from 38 countries with fieldwork from 1994 to 2024. We included 675,584 women aged 15–49 years with demand for family planning and grouped them into 144 intersectional strata by age, union status, education, household wealth and residence. We fitted multilevel logistic models with random effects for strata and surveys (multilevel analysis of individual heterogeneity and discriminatory accuracy, MAIHDA) and estimated the variance partition coefficient (VPC), the area under the curve (AUC) and the proportional change in variance (PCV). Weighted demand satisfied rose from 27.5% in 1994–99 to 54.1% in 2018–24. Predicted demand satisfied ranged from 13.2% (95% CI: 11.0 −15.9) among married adolescents with no schooling in the poorest rural households to 68.0% (95% CI: 63.6 - 72.0) among unmarried urban women aged 25–34 years with secondary schooling in the richest households. The strata explained only 6.9% (95% CI: 5.6 - 8.7) of individual variation, with small discriminatory accuracy (AUC = 0.65; 95% CI: 0.63 - 0.66), and a Bayesian model gave a similar VPC (7.0%; 95% credible interval: 5.7 - 8.9). Additive effects explained 95.1% of the between-stratum variance. The VPC fell from 10.1% (95% CI: 8.0 - 12.8) in 1994–99 to 4.5% (95% CI: 3.6 - 5.8) in 2018–24. Five strata of married rural women with no schooling were in the lowest decile in every period. In the latest surveys, married rural women aged 15–24 years with no schooling had lower demand satisfied than other women in 29 of 30 countries (median gap 17.5 percentage points). The 15 most disadvantaged strata held a median 16.4% of unsatisfied demand, so targeting these groups alone would reach about one in six women whose demand is not met. The main limitations are the cross-sectional design, self-reported contraceptive use and the exclusion of 15 early surveys.

**Conclusions:** Intersectional gaps in demand satisfied by modern methods have narrowed in sub-Saharan Africa, and most of the remaining gap between groups is additive. Social position alone discriminates poorly between women, so programmes targeted only by social group would miss most women with unmet need. This supports strengthening universal services. Within universal services, married adolescents and young women with no schooling in poor rural households have been left behind for three decades in almost every country and need priority, country-specific programmes if the 2030 targets are to be met.

**Author summary:** *Why was this study done?:*

- Sustainable Development Goal 3.7 commits countries to universal access to family planning by 2030, and the share of women whose demand is satisfied by modern methods is the main indicator of progress.
- Gaps in this indicator are usually reported by one characteristic at a time, such as age or wealth, although women live at the intersection of several characteristics.
- Programme managers need to know whether to target particular social groups or to strengthen services for all women, and which groups, if any, have been left behind.

*What did the researchers do and find?:*

- We analysed 675,584 women with demand for family planning in 139 national surveys from 38 sub-Saharan African countries between 1994 and 2024, grouped into 144 combinations of age, marital status, schooling, wealth and residence.
- Demand satisfied ranged from 13% to 68% across groups, but group membership explained only 7% of the differences between individual women, and the gaps between groups halved over three decades.
- Married adolescents and young women with no schooling in poor rural households were among the most disadvantaged groups in every period, and in the latest surveys they lagged behind other women in 29 of 30 countries by a median of 17.5 percentage points.

*What do these findings mean?:*

- Targeting family planning programmes only by social characteristics would miss most women whose demand is not met, so universal services should remain the foundation.
- Ministries of health and FP2030 partners should give priority to married adolescents and young women with no schooling in rural areas, and should report SDG indicator 3.7.1 for this group alongside the national average.
- The study describes associations from self-reported cross-sectional survey data and cannot identify the causes of these gaps.

## Introduction

The proportion of women whose demand for family planning is satisfied by modern methods is indicator 3.7.1 of the Sustainable Development Goals (SDGs). It is a core measure of progress towards universal access to sexual and reproductive health care [1–3]. A threshold of 75% has been proposed as the level that countries should reach by 2030 [2,4]. Sub-Saharan Africa has made the largest gains of any region since the 1990s. Yet it still has the lowest levels of demand satisfied, and progress has been uneven between and within countries [5–9]. The fertility transition in the region has also been slower than elsewhere [10,11].

Studies of who is left behind usually describe gaps along one dimension at a time, such as age, wealth, residence or marital status [12,13]. In multi-country analyses, adolescents, women with no schooling, poorer women and rural women have lower demand satisfied [12,14–16]. These characteristics do not occur in isolation. A woman is not only young or poor or rural. She may be all of them at once. Intersectionality theory holds that social positions combine to shape access to resources and services. The experience of people at particular intersections may therefore not be predicted by adding up single-axis disadvantages [17,18].

Single-dimension analyses cannot show whether gaps combine additively or multiply. Conventional regression with many interaction terms also becomes unstable when strata are numerous and some are small [19,20]. Multilevel analysis of individual heterogeneity and discriminatory accuracy (MAIHDA) addresses these problems [19,21,22]. Individuals are nested in intersectional strata, and the stratum random effects give shrunken and stable estimates for every stratum. The approach also measures how well the strata discriminate between individuals with and without the outcome, through the variance partition coefficient (VPC) and the area under the receiver operating characteristic curve (AUC) [23,24]. It separates additive effects from interaction effects [25]. MAIHDA has been used for chronic disease, mental health and, recently, child anaemia in Latin America [24,26]. We found no published study that has applied it to demand satisfied by modern methods or to other contraceptive outcomes in sub-Saharan Africa.

Knowing which combinations of characteristics leave women behind matters for programmes. If disadvantage is mostly additive, broad strategies that address each characteristic may be enough. If particular combinations fare worse than expected, programmes need to be designed for them. If social position discriminates poorly between women, targeting by social characteristics alone will miss many women in need. These questions have not been answered for family planning in sub-Saharan Africa.

We aimed to quantify intersectional inequalities in demand satisfied by modern methods among women in sub-Saharan Africa from 1994 to 2024. We estimated how demand satisfied varied across 144 strata defined by age, union status, education, household wealth and residence. We assessed how much of the individual variation lay between strata and how much of it was additive. We also examined how inequalities changed over time, how they differed between sub-regions and countries, and which groups were persistently left behind.

We preregistered five hypotheses. First, demand satisfied would be lowest among adolescent, unmarried, poorer, rural women with no schooling (H1). Second, the strata would have low to moderate discriminatory accuracy, with a VPC below 20% and an AUC below 0.70 (H2). Third, additive effects would explain more than 80% of the between-stratum variance, with some interaction effects (H3). Fourth, the between-stratum variance would be smaller in 2018–24 than in the 1990s, with the least improvement in strata combining adolescence, no schooling and rural residence (H4). Fifth, inequalities would be wider in west and central Africa than in east and southern Africa (H5).

## Methods

### Study design, data sources and ethics statement

This was a preregistered cross-sectional secondary analysis of Demographic and Health Surveys (DHS). The analysis plan was registered on OSF Registries on 19th October 2026, before any data were downloaded (doi:10.17605/OSF.IO/8M9GB; S2 Appendix). It is reported following the Strengthening the Reporting of Observational Studies in Epidemiology (STROBE) guideline (S1 Checklist) [27]. DHS are nationally representative household surveys with two-stage stratified cluster sampling. Trained interviewers administer standard questionnaires face to face [28]. We used the women’s individual recode files of all standard DHS done in sub-Saharan Africa with fieldwork from 1992 to 2024 for the 41 countries approved by the DHS Program for this project. Interim, malaria indicator, AIDS indicator and subnational surveys were excluded. For 30 older surveys in the main analysis whose women’s file did not include wealth quintiles, we merged the DHS wealth index file through the household identifier.

This analysis used publicly available, anonymised DHS data, so no ethical approval was needed. Access to the data was granted by the DHS Program (project 246219, 19th October 2026). Informed consent was obtained from all respondents during the original surveys, and the authors had no contact with participants.

Of 154 eligible surveys, 13 did not measure demand for family planning among unmarried women. These were the 12 DHS-II surveys and the 1994 survey in Côte d’Ivoire, which asked fertility preferences only of women in union [29]. As the protocol specifies for ever-married samples, these were used only in a sensitivity analysis of women in union. Mozambique 2003 and Tanzania 1999 were excluded because our national estimates differed from the published values by more than 1 percentage point and the difference could not be resolved. Chad 2004 and Niger 2006 also differed from the published values, but our estimates agreed with each survey’s own unmet-need variable, so we judged the published values to be in error and retained these two surveys. The main analysis therefore included 139 surveys from 38 countries with fieldwork from 1994 to 2024 (S1 Fig and S1 Appendix, Table S1). These surveys interviewed 1,697,188 women aged 15–49 years, of whom 675,897 had demand for family planning. We excluded 313 women (0.05%) with a missing stratification variable, which left 675,584 women.

### Outcome

Women with demand for family planning were those currently using contraception and those with unmet need for spacing or limiting, according to the revised DHS definition [29]. Under this definition, women not in union are classified as having unmet need only if they had sexual intercourse in the four weeks before the survey, whereas women in union are classified regardless of recent sexual activity. The outcome was coded 1 if a woman with demand was using a modern method and 0 otherwise [27]. We used demand satisfied rather than contraceptive prevalence or unmet need because it is the SDG indicator and it compares groups with different fertility intentions on the same scale [2]. The revised unmet-need variable is included in the recode files of most surveys from 2010 onwards. For the 66 earlier surveys that did not include it, we computed it with a Python translation of the DHS Program’s published Stata code, including its survey-specific rules. In 24 surveys that included the official variable, our classification agreed with it for 99.8% of women. Demand satisfied differed by 0.2 percentage points on average (S1 Appendix, Table S2). We also reproduced the national estimate of demand satisfied for each survey and compared it with the published STATcompiler value.

### Intersectional strata

Strata were defined by five preregistered dimensions. These were age (15–19, 20– 24, 25–34 and 35–49 years), union status (married or living with a partner, or not in union), education (none, primary, or secondary or higher), household wealth (poorest 40%, middle 20% or richest 40% of the survey’s wealth index [30]) and residence (urban or rural). Their combination gives 144 strata, all of which were represented. The smallest stratum had 140 women. These dimensions are those used to monitor equity in family planning and were fixed in the protocol [12]. Religion and ethnicity were not included because their categories are not comparable across countries. Parity was examined as a sixth dimension in a sensitivity analysis.

### Statistical analysis

Weighted descriptive estimates used the DHS sampling weights, rescaled so that each survey contributed equally. We followed the MAIHDA approach for the main models [19,21,22]. Model 1 (null) was a logistic model with random intercepts for intersectional strata and, cross-classified, for surveys. Model 2 (additive) added the main effects of the five dimensions. Model 3 added period (1994–99, 2000–09, 2010–17 and 2018–24). From Model 1 we calculated the VPC as the stratum variance divided by the sum of the stratum variance, the survey variance and π²/3. We calculated the AUC from predictions that included the fixed intercept and the stratum effects but not the survey effects, so that it measures the discriminatory accuracy of intersectional position alone. We also report the median odds ratio (MOR), which expresses the stratum variance on the odds ratio scale. It is the median odds ratio between a woman in a stratum with higher demand satisfied and a woman in a stratum with lower demand satisfied when two strata are chosen at random [23]. An AUC below 0.60 was interpreted as absent or poor discrimination, 0.60 to 0.70 as small, 0.70 to 0.80 as moderate and above 0.80 as large.

The proportional change in variance (PCV) was the reduction in stratum variance from Model 1 to Model 2. The stratum random effects in Model 2 were interpreted as interaction effects. They show how far a stratum departs from the prediction based on additive main effects. An interaction effect was considered present when its 95% interval excluded zero. Predicted probabilities for each stratum came from Model 1, with 95% intervals from 2000 simulations of the fixed intercept and the conditional distribution of the stratum effect.

Models 1 and 2 were refitted within each period, each sub-region and each country. Sub-regions were west and central Africa (WCA) and east and southern Africa (ESA), following the UNICEF grouping. Model 6 added random effects for stratum- by-period combinations to estimate stratum trajectories. A stratum caught up or fell further behind if its change in predicted demand satisfied from 1994–99 to 2018–24 differed from the average stratum change and the 95% simulation interval of the difference excluded zero. Model 7 added random effects for stratum-by-country combinations. A stratum was persistently left behind if it was in the lowest 10% of predicted demand satisfied in every period.

We checked whether the trends were driven by changes in which countries were surveyed. We refitted the period models in the eight countries with a survey in every period and in the 11 countries with surveys in both 1994–99 and 2018–24. To make the results usable by national programmes, we did an analysis that was not preregistered and compared weighted demand satisfied in each country’s latest survey between married rural women aged 15–24 years with no schooling and all other women with demand. This group combines the characteristics of the persistently left-behind strata. Confidence intervals used the effective sample size, and countries with fewer than 30 women in the group were not shown.

The outcome and all stratum-defining variables are categorical. The models were therefore fitted to binomial counts for each stratum within each survey, which gives the same likelihood as an individual-level model. Because the counts were aggregated to strata within surveys, survey clusters could not be included as an additional level. Models were estimated by maximum likelihood with the Laplace approximation in R version 4.3 using lme4 [31]. Equivalent Stata code using melogit is available from the corresponding author. Confidence intervals for the stratum variance and VPC were obtained by profile likelihood. Confidence intervals for the pooled VPC and AUC were also estimated by a cluster bootstrap in which surveys were resampled with replacement (200 replicates). At country level, we examined whether between-stratum inequality was lower in countries with higher overall demand satisfied, using Spearman correlation. Main multilevel models were unweighted, as preregistered, because weighting can bias variance components [32].

### Sensitivity and exploratory analyses

Preregistered sensitivity analyses used wealth quintiles (240 strata) and parity as a sixth dimension (432 possible strata, 431 observed). We restricted the analysis to women in union, including eight of the 13 surveys that did not measure unmet need among unmarried women. The other five have no wealth index. We also fitted models weighted with scaled survey weights, excluded surveys with fieldwork in 2020–21, excluded strata with fewer than 30 women and fitted Bayesian models in brms with weakly informative priors [33]. These were a normal(0, 2.5) prior for the intercept and half-Student t(3, 0, 1) priors for the stratum and survey standard deviations, with four chains of 2,500 iterations including 1,000 warm-up iterations. R-hat was 1.02 for the stratum and survey standard deviations. Mixing of the intercept was slower (R-hat 1.09), which does not affect the VPC (S1 Appendix). We additionally excluded Chad 2004 and Niger 2006, whose published values differed from ours (see Results). Multiple imputation was not needed because no survey had more than 5% of women excluded. Surveys are nested in countries, and repeated surveys from one country are not independent. We therefore fitted a three-level sensitivity model, which was not preregistered, in which strata were crossed with surveys nested in countries. This model separates stable differences between countries from differences between surveys within a country. We compared its VPC, PCV and interaction effects with those of the main model and tested the country variance with a likelihood ratio test.

In an exploratory analysis that was not preregistered, we examined whether factors that may lie on the pathway between social position and contraceptive use explained between-stratum differences. These were parity, weekly exposure to mass media, hearing family planning messages, a visit from a family planning worker, being told about family planning at a health facility and current employment. This analysis used the 105 surveys in which these variables were available for more than 90% of women. Deviations from the protocol are listed in S1 Appendix (section S3). To show what the stratum gaps mean for national coverage, we did a second exploratory analysis in the latest survey of each country. We defined the left-behind strata as the 15 strata in the lowest 10% of predicted demand satisfied in 2018–24. For each country we estimated their weighted share of all women with demand and their share of all women whose demand was not satisfied by modern methods. The ratio of these shares shows whether unsatisfied demand is concentrated in these strata. We also estimated how much national demand satisfied would rise if women in these strata had the same level as other women in that country. We repeated this for married rural women aged 15–24 years with no schooling.

## Results

The 139 surveys included 675,584 women with demand for family planning. There were 21 surveys (57,859 women) from 1994–99, 42 (166,848) from 2000–09, 46 (254,335) from 2010–17 and 30 (196,542) from 2018–24 (Table 1). Of these women, 80.2% were in union, 32.5% had no schooling and 60.3% lived in rural areas (weighted). National estimates were within 1 percentage point of the published value in 133 of the 137 main surveys with a published value. Rwanda 2010 and Uganda 2011 matched the published value for women in union exactly. For Chad 2004 and Niger 2006, our estimates were close to those from each survey’s own unmet-need variable based on the original definition (6.3% and 19.2%). They were also consistent with the low modern contraceptive prevalence among married women in these surveys (1.6% and 5.0%). We therefore retained them, and excluding them did not change the results (S1 Appendix, Tables S1 and S6).

**Table 1.** Characteristics of women with demand for family planning and weighted demand satisfied by modern methods, 139 surveys in 38 sub-Saharan African countries, 1994 to 2024 (n = 675,584)

| Characteristic | Women with demand, n (weighted %) | Demand satisfied, weighted % | 1994–99 | 2000–09 | 2010–17 | 2018–24 |
|---|---|---|---|---|---|---|
| Age, years |  |  |  |  |  |  |
| 15–19 | 72,578 (10.6) | 35.0 | 20.5 | 31.5 | 39.5 | 46.5 |
| 20–24 | 134,523 (19.9) | 42.3 | 27.7 | 36.3 | 47.0 | 55.2 |
| 25–34 | 256,756 (38.6) | 44.1 | 28.8 | 37.5 | 48.6 | 56.4 |
| 35–49 | 211,727 (30.9) | 41.8 | 28.6 | 34.6 | 46.4 | 53.0 |
| Union status |  |  |  |  |  |  |
| In union | 534,423 (80.2) | 39.1 | 24.7 | 32.2 | 43.5 | 52.4 |
| Not in union | 141,161 (19.8) | 54.3 | 39.2 | 50.7 | 60.0 | 60.1 |
| Education |  |  |  |  |  |  |
| No education | 194,626 (32.5) | 26.0 | 11.9 | 16.9 | 31.4 | 43.5 |
| Primary | 242,268 (34.4) | 44.2 | 29.3 | 37.2 | 49.8 | 59.9 |
| Secondary or higher | 238,690 (33.1) | 55.6 | 49.3 | 52.7 | 58.8 | 57.4 |
| Household wealth |  |  |  |  |  |  |
| Poorest 40% | 242,928 (33.9) | 32.7 | 17.1 | 24.5 | 38.0 | 46.6 |
| Middle 20% | 131,042 (19.1) | 40.0 | 23.8 | 31.3 | 45.1 | 53.9 |
| Richest 40% | 301,614 (47.0) | 49.7 | 36.0 | 45.4 | 53.8 | 59.6 |
| Residence |  |  |  |  |  |  |
| Urban | 263,255 (39.7) | 50.0 | 39.6 | 46.0 | 53.4 | 55.4 |
| Rural | 412,329 (60.3) | 36.9 | 20.4 | 30.1 | 42.1 | 52.9 |
| Parity |  |  |  |  |  |  |
| 0 | 74,143 (10.9) | 45.0 | 29.2 | 42.4 | 50.2 | 52.3 |
| 1–2 | 212,659 (31.3) | 47.2 | 32.6 | 40.4 | 51.2 | 58.9 |
| 3 or more | 388,782 (57.8) | 38.7 | 24.7 | 32.1 | 43.6 | 51.5 |
| Sub-region |  |  |  |  |  |  |
| West and Central Africa | 327,983 (52.5) | 32.1 | 16.0 | 24.9 | 36.2 | 44.6 |
| East and Southern Africa | 347,601 (47.5) | 53.1 | 40.1 | 45.6 | 58.2 | 68.4 |
| Total | 675,584 (100) | 42.1 | 27.5 | 35.7 | 46.7 | 54.1 |

Weighted percentages use DHS sampling weights rescaled so that each survey contributes equally. The sub-region distribution therefore reflects the number of surveys (73 and 66). Numbers of surveys by period were 21, 42, 46 and 30.

Weighted demand satisfied by modern methods increased from 27.5% in 1994–99 to 46.7% in 2010–17 and 54.1% in 2018–24 (Table 1). It was lowest among adolescents aged 15–19 years (35.0%), women with no schooling (26.0%), women in the poorest 40% (32.7%) and rural women (36.9%). It was higher among women not in union (54.3%) than among women in union (39.1%). The gap between women with no schooling and those with secondary schooling narrowed from 37.3 percentage points in 1994–99 to 13.9 points in 2018–24.

Predicted demand satisfied varied more than five-fold across the 144 strata (Figure 1). It ranged from 13.2% (95% CI: 11.0 - 15.9) among married adolescents aged 15–19 years with no schooling in the poorest rural households to 68.0% (95% CI: 63.6 - 72.0) among unmarried women aged 25–34 years with secondary or higher schooling in the richest urban households. Of the 20 lowest strata, 18 were in union and 16 had no schooling. All 20 highest strata were women not in union, and 14 of them were urban.

**Figure 1.**
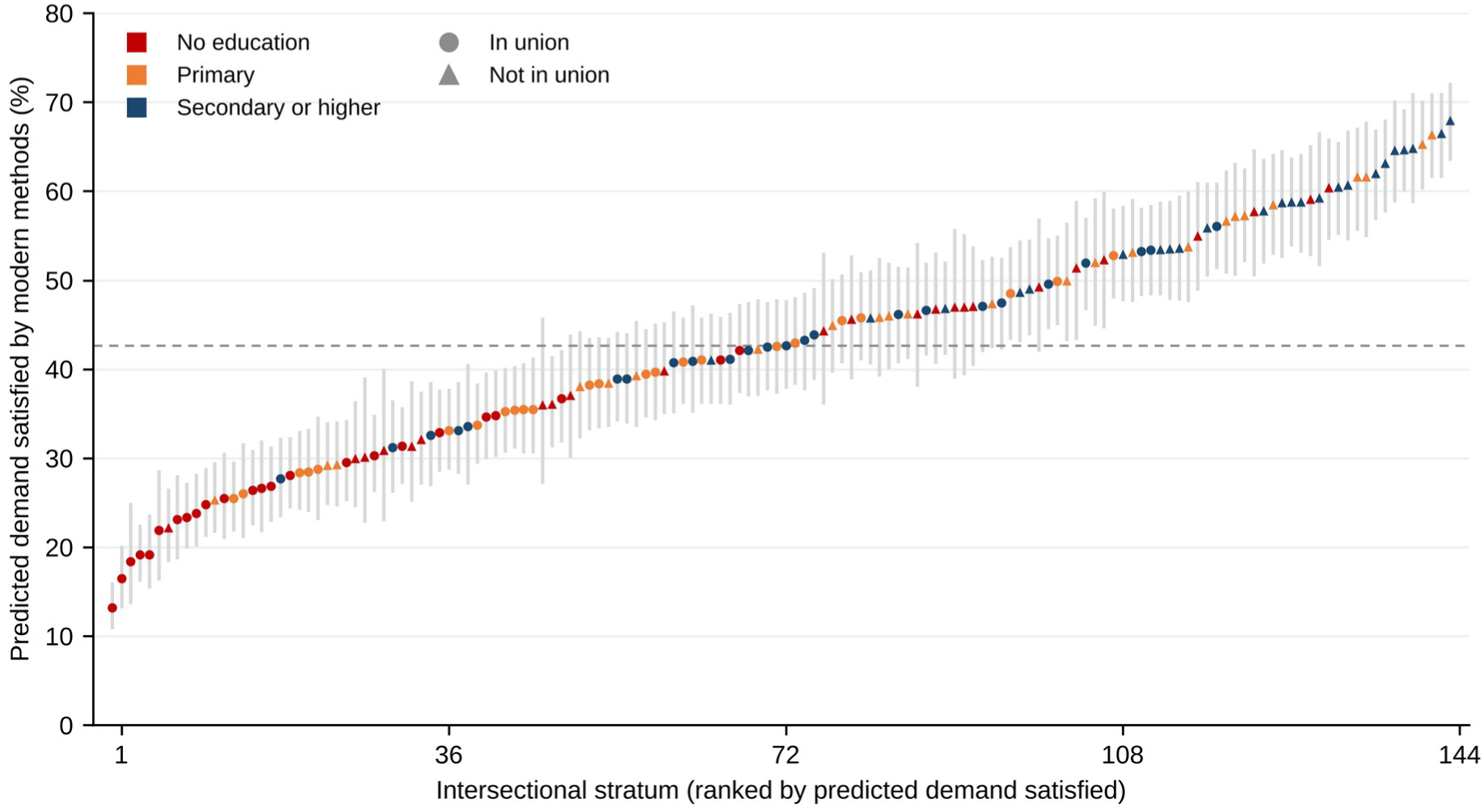
Predicted demand satisfied by modern methods in 144 intersectional strata, all surveys combined. Strata are ranked by predicted demand satisfied from the null model. Vertical lines show 95% intervals. Colour shows education and marker shape shows union status. The dashed line is the median of strata.

In the null model, 6.9% (95% CI: 5.6 - 8.7) of the individual variation in the latent propensity to have demand satisfied lay between strata. The strata had small discriminatory accuracy (AUC = 0.65) (Table 2). The cluster bootstrap gave a 95% CI of 6.1 - 8.0 for the VPC and 0.633 - 0.662 for the AUC. The MOR was 1.70 for strata and 2.40 for surveys. Survey differences accounted for more variation than strata. A model with survey effects alone gave an AUC of 0.72, which rose to 0.76 when stratum effects were added. In the additive model, demand satisfied was higher among women aged 25–34 years (aOR = 1.94; 95% CI: 1.82 - 2.07) than among adolescents. It was also higher among women with primary schooling (aOR = 1.48; 95% CI: 1.40 - 1.57) or secondary or higher schooling (aOR = 1.96; 95% CI: 1.85 - 2.07), women in the richest 40% (aOR = 1.58; 95% CI: 1.50 - 1.67) and urban women (aOR = 1.29; 95% CI: 1.23 - 1.35). It was lower among women in union than among unmarried women (aOR = 0.56; 95% CI: 0.53 - 0.58) (S1 Appendix, Table S8).

**Table 2.** Variance components, discriminatory accuracy and explained variance of intersectional strata, by period and sub-region.

|  | **Surveys** | **Women** | **Stratum variance (null)** | **Survey variance (null)** | **VPC, % (95% CI)** | **MOR** | **Stratum variance (additive)** | **PCV, %** | **AUC** |
|---|---|---|---|---|---|---|---|---|---|
| All surveys | 139 | 675,584 | 0.306 | 0.839 | 6.9 (5.6 - 8.7) | 1.70 | 0.015 | 95.1 | 0.65 |
| 1994–99 | 21 | 57,859 | 0.461 | 0.813 | 10.1 (8.0 - 12.8) | 1.91 | 0.025 | 94.6 | 0.72 |
| 2000–09 | 42 | 166,848 | 0.427 | 0.546 | 10.0 (8.1 - 12.5) | 1.86 | 0.013 | 97.0 | 0.69 |
| 2010–17 | 46 | 254,335 | 0.352 | 0.584 | 8.3 (6.7 - 10.4) | 1.76 | 0.020 | 94.3 | 0.65 |
| 2018–24 | 30 | 196,542 | 0.198 | 0.880 | 4.5 (3.6 - 5.8) | 1.53 | 0.014 | 92.7 | 0.60 |
| West and central Africa | 73 | 327,983 | 0.370 | 0.459 | 9.0 (7.3 - 11.2) | 1.79 | 0.010 | 97.4 | 0.65 |
| East and southern Africa | 66 | 347,601 | 0.276 | 0.847 | 6.3 (5.0 - 7.9) | 1.65 | 0.025 | 91.0 | 0.64 |
| WCA 1994–99 | 11 | 25,120 | 0.585 | 0.209 | 14.3 | 2.07 | 0.033 | 94.4 | 0.73 |
| WCA 2000–09 | 20 | 74,549 | 0.509 | 0.220 | 12.7 | 1.98 | 0.011 | 97.8 | 0.70 |
| WCA 2010–17 | 24 | 117,644 | 0.496 | 0.263 | 12.3 | 1.96 | 0.020 | 96.0 | 0.66 |
| WCA 2018–24 | 18 | 110,670 | 0.224 | 0.285 | 5.9 | 1.57 | 0.004 | 98.3 | 0.60 |
| ESA 1994–99 | 10 | 32,739 | 0.425 | 0.804 | 9.4 | 1.86 | 0.018 | 95.8 | 0.70 |
| ESA 2000–09 | 22 | 92,299 | 0.351 | 0.469 | 8.5 | 1.76 | 0.016 | 95.3 | 0.68 |
| ESA 2010–17 | 22 | 136,691 | 0.265 | 0.527 | 6.5 | 1.63 | 0.030 | 88.7 | 0.63 |
| ESA 2018–24 | 12 | 85,872 | 0.216 | 1.054 | 4.7 | 1.56 | 0.038 | 82.6 | 0.60 |

Variances are on the log-odds scale. VPC is the stratum variance divided by the sum of the stratum variance, the survey variance and π²/3. PCV is the proportional change in stratum variance from the null to the additive model. AUC is for predictions including stratum effects. 95% CIs for the VPC are from the profile likelihood of the stratum variance. MOR is the median odds ratio for strata, exp(√(2 × stratum variance) × 0.6745).

The additive effects explained 95.1% of the between-stratum variance. Nevertheless, 59 strata had interaction effects whose 95% intervals excluded zero. Of these, 31 had higher and 28 had lower demand satisfied than predicted by the additive effects (S1 Appendix, Table S4 and S2 Fig). The largest negative interaction was among unmarried rural adolescents with primary schooling in the richest households (−10.8 percentage points). Unmarried rural adolescents with primary schooling had negative interactions in every wealth group. Because stratum effects are shrunk towards zero, these interaction estimates are conservative.

Between-stratum inequality fell over time (Figure 2 and Table 2). The VPC was 10.1% (95% CI: 8.0 - 12.8) in 1994–99, 10.0% in 2000–09, 8.3% in 2010–17 and 4.5% (95% CI: 3.6 - 5.8) in 2018–24. Over the same period the AUC fell from 0.72 to 0.60, and the stratum variance fell from 0.46 to 0.20 on the log-odds scale. The decline was not explained by changes in which countries were surveyed. In the eight countries surveyed in every period, the VPC fell from 13.5% (95% CI: 10.5 - 17.5) to 4.6% (95% CI: 3.5 - 6.1). The AUC in these countries fell from 0.71 to 0.61 (S1 Appendix, Table S9).

**Figure 2.**
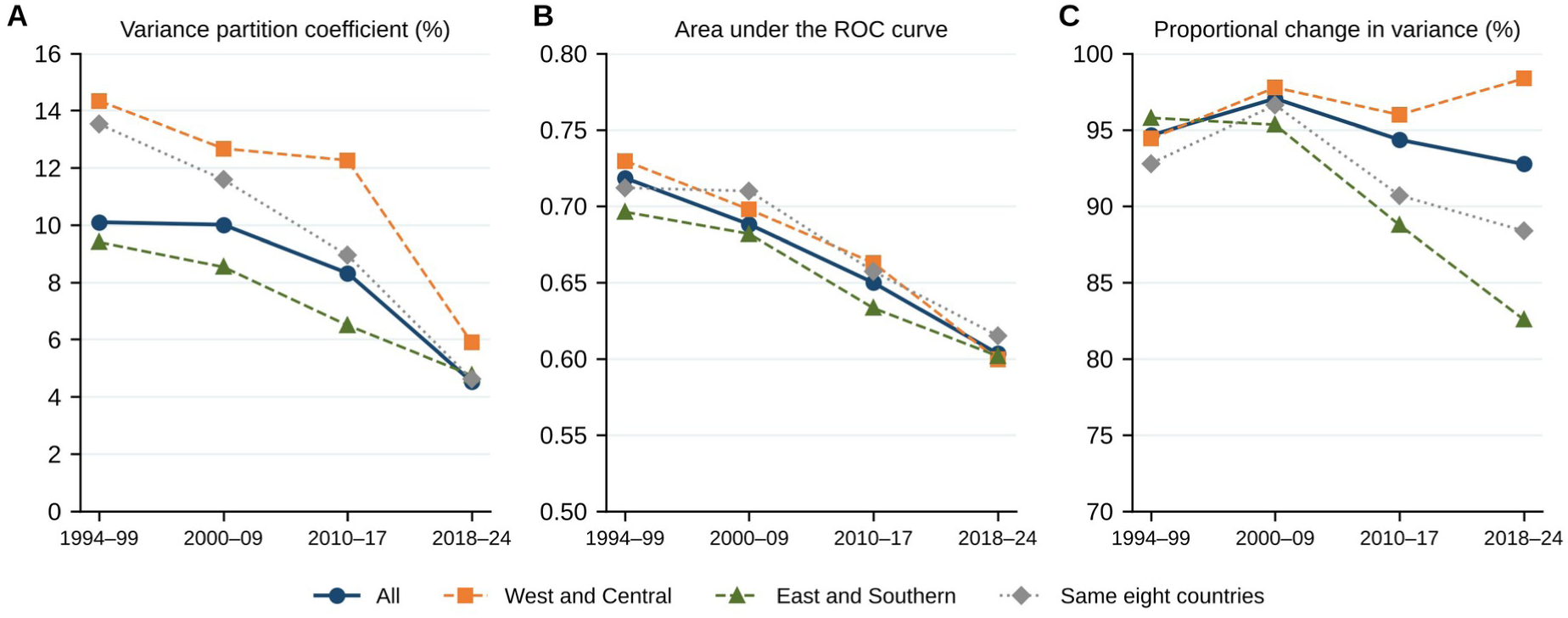
Discriminatory accuracy of intersectional strata by period and sub-region. (A) Variance partition coefficient. (B) Area under the receiver operating characteristic curve. (C) Proportional change in variance. The dotted line shows the eight countries surveyed in every period.

Inequality was somewhat higher in west and central Africa (VPC = 9.0%; 95% CI: 7.3 - 11.2) than in east and southern Africa (VPC = 6.3%; 95% CI: 5.0 - 7.9). The confidence intervals overlapped slightly. Unweighted demand satisfied was higher in east and southern Africa (57.2%) than in west and central Africa (33.7%). In west and central Africa, the VPC stayed above 12% until 2010–17 and then fell to 5.9%. In east and southern Africa, the share of stratum variance explained by additive effects fell from 95.8% to 82.6%. Interactions therefore accounted for a larger share of the smaller remaining inequality.

Across strata, predicted demand satisfied increased by an average of 27.1 percentage points between 1994–99 and 2018–24 (Figure 3). Thirteen strata caught up and gained 6 to 11 points more than average. All were rural, and ten had primary schooling. Fourteen strata gained less than average. Eleven of these were in the richest 40% and had started from relatively high levels. Most were urban women with secondary schooling, which is consistent with convergence rather than falling behind. The other three had started among the lowest and fell further behind. They were adolescents with no schooling in the poorest households. These were married adolescents in rural areas (Model 6 prediction from 6.7% to 24.5%), married adolescents in urban areas and unmarried adolescents in rural areas.

**Figure 3.**
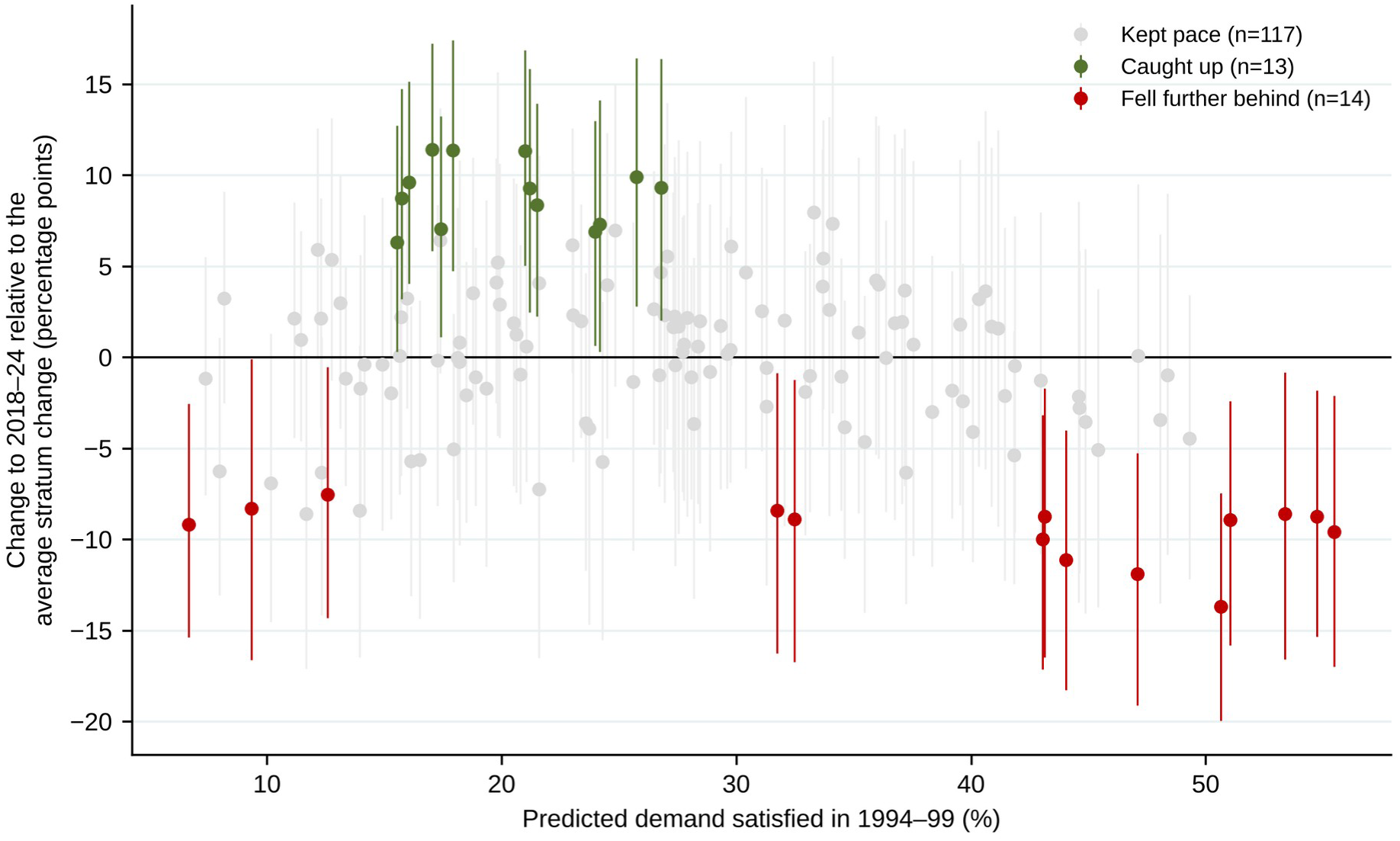
Change in predicted demand satisfied between 1994–99 and 2018–24 for each stratum. Change is shown relative to the average change across strata (27.1 percentage points) and plotted against the predicted level in 1994–99. Vertical lines show 95% simulation intervals. Strata were classified as having caught up or fallen further behind when the interval excluded zero.

Five strata were in the lowest decile in every period (Table 3). All were married rural women with no schooling, and four were aged 15–24 years.

**Table 3.** Strata persistently left behind. Predicted demand satisfied by modern methods (%) in strata that were in the lowest 10% of the 144 strata in every period.

| **Stratum** | **1994–99** | **2000–09** | **2010–17** | **2018–24** |
|---|---|---|---|---|
| 15–19, in union, no education, poorest 40%, rural | 7.0 | 8.5 | 14.8 | 25.9 |
| 15–19, in union, no education, middle 20%, rural | 7.7 | 10.1 | 20.9 | 33.2 |
| 20–24, in union, no education, poorest 40%, rural | 6.5 | 12.3 | 22.7 | 34.0 |
| 15–19, in union, no education, richest 40%, rural | 9.9 | 15.3 | 23.7 | 34.8 |
| 25–34, in union, no education, poorest 40%, rural | 7.7 | 15.3 | 27.7 | 38.8 |
| Median of all 144 strata | 27.2 | 37.0 | 48.7 | 56.8 |

Predictions are from period-specific null models with stratum and survey random intercepts.

In Model 7, the stratum-by-country variance (0.113) was much larger than the residual stratum variance common to all countries after additive and period effects (0.004). The pattern of intersectional inequality therefore differed substantially between countries. In country-specific models, the VPC ranged from 2.6% in Ghana to 29.1% in Chad. It exceeded 10% in 19 of 38 countries, most of which were in west and central Africa (S1 Appendix, Table S5 and S3 Fig). The country AUC ranged from 0.59 to 0.78. Countries with higher overall demand satisfied had lower between-stratum inequality (Spearman ρ = -0.47; p = 0.003).

In the latest survey of each country, married women aged 15–24 years with no schooling in rural areas had lower demand satisfied than other women in 29 of the 30 countries with at least 30 such women (Figure 4). The 95% CI excluded the value for other women in 25 countries. The median gap was 17.5 percentage points, and the gap was 20 points or more in 12 countries. It was largest in Kenya (45 points), Sierra Leone (32 points) and Mozambique (32 points). In these countries, demand satisfied among other women was above 50%. Only Burundi had a higher level in this group, and the gap was small in Malawi (S1 Appendix, Table S10). The size of the gap was not related to the level of demand satisfied among other women (Spearman ρ = -0.19; p = 0.32).

**Figure 4.**
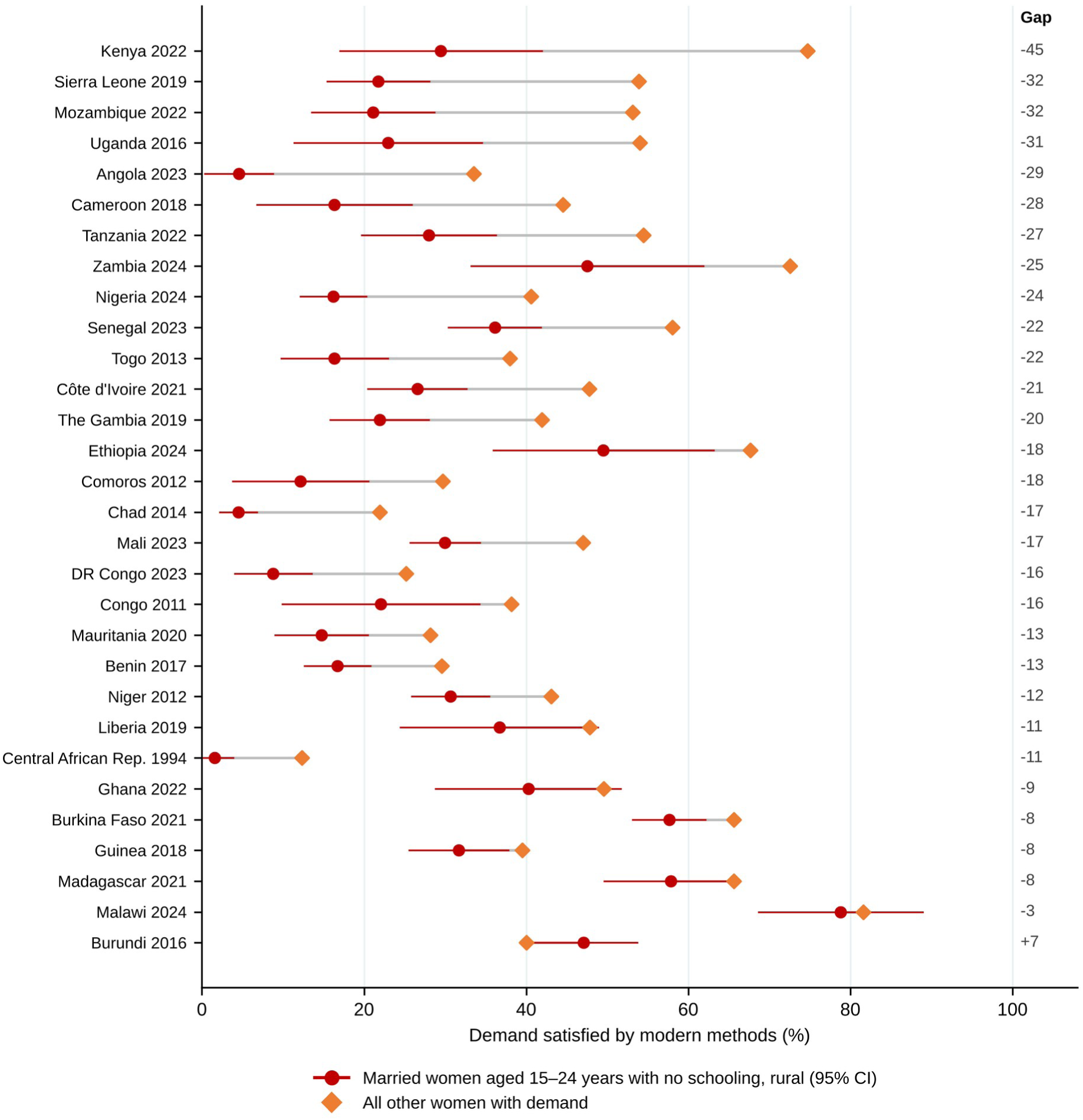
Demand satisfied among married rural women aged 15–24 years with no schooling compared with all other women, by country. Weighted estimates from the latest survey of each country with at least 30 women in the group. Horizontal lines show 95% CIs. Gap is the difference in percentage points.

Unsatisfied demand was concentrated in the left-behind strata. In the median country, these 15 strata included 11.7% of women with demand but 16.4% of women whose demand was not satisfied by modern methods. Their share of unsatisfied demand was larger than their share of demand in all 38 countries (median ratio 1.33). It was highest in Guinea (38.5%). If women in these strata had the same level as other women, national demand satisfied would rise by a median of 1.5 percentage points, and by up to 6.5 points in Guinea. Married rural women aged 15– 24 years with no schooling made up 3.0% of women with demand and 4.0% of those with unsatisfied demand in the median country. Their share of unsatisfied demand was larger than their share of demand in 29 of 30 countries. In every country, most women with unsatisfied demand were outside the left-behind strata (S1 Appendix, Table S11).

Results were robust in sensitivity analyses (S1 Appendix, Table S6). The VPC ranged from 4.7% to 8.8%, the AUC from 0.64 to 0.65 and the PCV from 89.1% to 97.0%. Adding parity as a sixth dimension increased the VPC to 8.8%. Restriction to women in union reduced it to 4.7%. The Bayesian null model gave a posterior median VPC of 7.0% (95% CrI: 5.7 - 8.9). The Bayesian PCV was 94.8% (95% CrI: 92.4 - 96.4). In the three-level model, the stratum VPC was 6.9% (95% CI: 5.6 - 8.6), the same as in the main model. Countries accounted for 9.0% of the variation and surveys within countries for 10.3%. The country variance was significant (likelihood ratio χ² = 31.7; p < 0.001), and countries explained 47% of the between-survey variance. The PCV was 95.1%, and the same 59 strata had interaction effects. Stratum effects from the two models were almost identical (r > 0.99) (S1 Appendix, Table S6).

In the exploratory analysis of 105 surveys, being told about family planning at a health facility (aOR = 1.47; 95% CI: 1.44 - 1.49), employment (aOR = 1.27; 95% CI: 1.26 - 1.29), hearing family planning messages (aOR = 1.23; 95% CI: 1.22 - 1.25), weekly media exposure (aOR = 1.17; 95% CI: 1.15 - 1.19) and a visit from a family planning worker (aOR = 1.13; 95% CI: 1.11 - 1.15) were associated with demand satisfied (S1 Appendix, Table S7). Without the stratum main effects but with period, these factors explained 12.1% of the between-stratum variance. Added to the additive model, they increased the explained share only from 95.1% to 95.2%.

## Discussion

In this preregistered analysis of 675,584 women with demand for family planning in 38 countries over three decades, demand satisfied by modern methods varied five-fold across intersectional strata. Intersectional position, however, explained only about 7% of the individual variation and had small discriminatory accuracy. Most of the between-stratum inequality was explained by the additive effects of age, schooling, wealth, residence and union status. This inequality halved between 1994–99 and 2018–24, and the decline was the same in countries surveyed in every period. These broad gains hide groups that have not benefited. Married adolescents and young women with no schooling in poor rural households were among the lowest-ranked groups in every period. In the latest surveys, this group lagged behind other women in almost every country.

These findings support four of our five preregistered hypotheses. The strata had small discriminatory accuracy (H2). Additive effects explained most of the between-stratum variance, alongside some interaction effects (H3). Between-stratum variance fell over time, while the poorest adolescents with no schooling improved least (H4). Inequality was higher in west and central Africa than in east and southern Africa, although the difference was modest and the confidence intervals overlapped, so H5 was only partly supported. H1 was supported for age, schooling, wealth and residence. Contrary to our hypothesis, women in union, not unmarried women, had lower demand satisfied.

The narrowing of inequality is consistent with the inverse equity hypothesis. It predicts that new interventions first reach the better off and that gaps narrow once better-off groups approach saturation [34,35]. Strata that started high, mostly urban women with secondary schooling in the richest households, gained less than average. Rural women with primary schooling gained most. The same pattern has been described for institutional delivery across hundreds of national surveys [35]. At country level, inequality between strata was lower where overall demand satisfied was higher, which fits the same pattern. These data are consistent with the hypothesis but cannot prove it. The strata that started lowest did not all follow this pattern. Married adolescents and young women with no schooling in poor rural households stayed among the lowest-ranked strata in every period. This suggests that general expansion of services does not resolve the mechanisms that exclude them.

Married adolescents face specific barriers. These include provider bias, lack of privacy, pressure to prove fertility early in marriage and limited decision-making power within the household [15,16]. Exposure to family planning information is also lower among married adolescents in west Africa [36]. Programmes designed for this group can work. In a cluster randomised trial in Niger, a gender-synchronised programme for married adolescent girls and their husbands increased modern contraceptive use [37]. Our results suggest that such programmes are needed in almost every country in the region, not only in countries with low overall coverage.

The low discriminatory accuracy of the strata is an important result for policy. A small VPC does not mean that the gaps between groups are unimportant. Predicted demand satisfied still varied five-fold between strata. It means that women within the same stratum differ greatly from each other. An AUC of 0.65 means that knowing a woman’s age, union status, schooling, wealth and residence gives only a limited indication of whether her demand is met. Interventions targeted only by these characteristics would therefore miss many women in need while also reaching many who are already served [21,23]. This supports universal provision of services, combined with proportionately greater effort for the most disadvantaged groups. Survey context had more discriminatory value than intersectional position. This points to the importance of national policy, commodity supply and health system factors [11,38].

The large stratum-by-country variance shows that the same social position carries different risks in different countries. National programmes therefore need to identify their own left-behind groups rather than rely on regional averages. Our country estimates show that young married rural women with no schooling lag behind in almost every country. This includes countries with high overall coverage such as Kenya, Sierra Leone and Mozambique, where demand satisfied among other women was above 50% and the gap was 32 to 45 points. The gap for this group was no smaller in countries with higher overall coverage, which suggests that general expansion of services alone does not close it. Reporting SDG indicator 3.7.1 only as a national average, or by one characteristic at a time, would miss these gaps. Our impact analysis adds a further point. The left-behind strata hold more than their share of unsatisfied demand, yet most women with unsatisfied demand belong to other strata. Closing the gap for the left-behind strata alone would raise national coverage by a median of only 1.5 percentage points. Targeted programmes for these groups are therefore needed for equity, but they cannot replace the expansion of services for all women. This supports an approach in which services stay universal and their intensity rises with disadvantage.

Women in union had lower demand satisfied than women not in union with the same age, schooling, wealth and residence. Unmarried women count as having demand only if they are sexually active, so they are a selected group, and many unmarried users rely on condoms and other short-acting methods [8,13]. For married women, partner and family expectations about fertility may limit use [39]. The restriction to women in union removes this selection, and in that analysis the VPC was 4.7%, while the AUC and the share of variance explained by additive effects were similar to the main analysis. This finding highlights married adolescents as a group in particular need. At the same time, the negative interaction effects among unmarried rural adolescents show that these adolescents face barriers that are not captured by the main effects.

The exploratory analysis showed that contact with family planning services, exposure to family planning messages and employment were associated with demand satisfied. These factors, however, explained little of the difference between strata beyond the additive effects. The disadvantage of the left-behind groups is therefore not simply due to less contact with services. It may reflect structural and normative barriers, such as partner opposition and early marriage, that our survey variables did not capture.

### Strengths and limitations

The study has several strengths. The analysis was preregistered and used standard surveys from almost all countries in the region over three decades. We applied one definition of unmet need to every survey, rebuilt the indicator for older surveys with the DHS Program’s own algorithm and validated it. We also checked every national estimate against the published value. MAIHDA gives stable estimates for small strata and separates additive from interaction effects. Results were consistent across preregistered sensitivity analyses, including Bayesian estimation, and in countries surveyed in every period. A three-level model that allowed for the nesting of surveys within countries gave the same results.

There are also limitations. First, the analysis is descriptive and cross-sectional, and the strata are markers of social position rather than causes. Second, the surveys included differ between periods, although the decline in between-stratum inequality was the same in the eight countries surveyed in every period. Third, contraceptive use, union status and sexual activity are self-reported. Use may be under-reported among unmarried women, which could bias their estimates in either direction. Fourth, the wealth index is relative within each survey, so a wealth group does not represent the same living standard across countries or periods. Fifth, the 13 surveys that did not measure unmet need among unmarried women and two surveys with unresolved differences from published estimates were excluded from the main analysis. This reduced coverage of the early 1990s, although the analysis of women in union that included eight of them gave similar results. Sixth, the interaction effects are descriptive and may partly reflect differences in composition within strata, such as parity or region. Seventh, the main models were unweighted and did not include survey clusters as a level, although models weighted with scaled survey weights gave similar results. As a result, differences between communities within a survey were not separated from individual variation. The categories of each dimension were also broad, and finer categories could reveal larger gaps. Finally, the exploratory variables were not available for all surveys and could not capture partner opposition, provider behaviour or service quality.

## Conclusions

Intersectional inequalities in demand satisfied by modern methods have narrowed in sub-Saharan Africa, and most of the remaining gaps between groups are additive. Social position alone discriminates poorly between women, which supports universal services. Married adolescents and young women with no schooling in poor rural households have, however, been left behind for three decades in almost every country. Reaching universal access by 2030 is likely to require universal services combined with country-specific programmes that reach these women. Countries should report SDG indicator 3.7.1 by combinations of social characteristics rather than one dimension at a time.

## Data Availability

No new data were generated by this study. The following existing data sources were used: the Demographic and Health Surveys (DHS) for the sub-Saharan African countries and survey years included in the analysis, available from the DHS Program at https://dhsprogram.com/data/available-datasets.cfm. The data are third-party data owned by the DHS Program and can be obtained free of charge by any researcher who registers and submits a brief project description at https://dhsprogram.com/data/new-user-registration.cfm. The authors accessed the data under DHS project number 246219 and had no special access privileges that others would not have. The Stata code used for data preparation and the multilevel analysis of individual heterogeneity and discriminatory accuracy (MAIHDA) is available on the Open Science Framework at https://osf.io/8m9gb.

https://dhsprogram.com/data/available-datasets.cfm

## Acknowledgments

We thank the DHS Program for access to the data and the women who took part in the surveys. The authors used an AI assistant (Claude, Anthropic) for language editing and formatting of the manuscript. The authors designed the study, conducted and verified all analyses, interpreted the findings and take full responsibility for the content.

## Supporting information

S1 Appendix. Supplementary tables S1 to S11 and methods. Surveys and national validation, validation of the computed unmet-need indicator, deviations from the protocol, results for all 144 strata, country results, sensitivity analyses, exploratory models, fixed effects, trends in countries surveyed in every period, country estimates for the left-behind group and the share of unsatisfied demand in the left-behind strata.

S2 Appendix. Preregistered analysis plan (OSF Registries, osf.io/8m9gb, 19th October 2026).

S1 Fig. Selection of surveys and women.

S2 Fig. Interaction effects for 144 intersectional strata.

S3 Fig. Variance partition coefficient and AUC by country.

S1 Checklist. STROBE checklist for cross-sectional studies.

## Author contributions (CRediT)

Conceptualization, PK. Data curation, PK. Formal analysis, PK. Methodology, PK. Project administration, PK. Supervision, PK. Validation, Visualization, PK. Writing (original draft), PK. Writing (review and editing), PK, SM, MN, AN-H.

## Data availability

The data are third-party data from the Demographic and Health Surveys Program and cannot be shared by the authors. They are available free of charge to registered researchers who request access at https://dhsprogram.com/data/. The authors had no special access privileges. All analysis code, the code that computes the revised unmet-need indicator and the list of survey files used are available from the corresponding author on reasonable request. The study protocol is registered at the Open Science Framework (osf.io/8m9gb) and provided in S2 Appendix.

## Abbreviations

aOR: adjusted odds ratio
AUC: area under the receiver operating characteristic curve
CI: confidence interval
CrI: credible interval
DHS: Demographic and Health Surveys
ESA: east and southern Africa
MAIHDA: multilevel analysis of individual heterogeneity and discriminatory accuracy
MOR: median odds ratio
PCV: proportional change in variance
SDG: Sustainable Development Goal
VPC: variance partition coefficient
WCA: west and central Africa

